# Non-temporal tree-based models outperform temporal deep learning models in the prediction of chemotherapy-induced side effects from longitudinal laboratory data

**DOI:** 10.64898/2025.12.12.25342142

**Authors:** Farnaz Rahimi, Christel Sirocchi, Julian Matschinske, Markus Metzler, Jakob Zierk, David B. Blumenthal

**Affiliations:** Biomedical Network Science Lab, Department Artificial Intelligence in Biomedical Engineering, Friedrich-Alexander-Universität Erlangen-Nürnberg, Nürnberger Str. 74, 91052 Erlangen, Germany.; Bitspark GmbH, Research and Development, Gebersdorfer Str. 277, 90449 Nürnberg, Germany.; Department of Pediatrics and Adolescent Medicine, University Hospital Erlangen, Loschgestr. 15, 91054 Erlangen, Germany.

**Author notes:** Contributing authors. These authors contributed equally to this work (Farnaz Rahimi, Christel Sirocchi).

**Keywords:** electronic health records, machine learning, prediction of chemotherapy-induced side effects, benchmark

## Abstract

The increasing availability of electronic health records (EHRs) provides opportunities to apply machine learning (ML) methods in support of clinical decision-making. The temporal nature of laboratory values in EHR data records makes them particularly suitable for temporal deep learning (DL) architectures that model patient trajectories over time. However, despite this potential, the application of temporal DL models to longitudinal laboratory data has largely been limited to intensive care unit (ICU) settings and coarse outcome prediction tasks such as mortality and readmission. How well they perform in sparse, irregular, and highly imbalanced data settings that are typical of clinical care outside of the ICU has not been fully assessed. To close this knowledge gap, we focused on the clinically important yet underexplored tasks to predict the chemotherapy-related complications aplasia and neutropenic fever before clinical onset, using longitudinal laboratory data extracted from EHR records from two independent datasets. Based on these datasets and targets, we systematically evaluated 13 ML models, including 7 state-of-the-art temporal DL models and 4 non-temporal tree-based baselines. Across all combinations of datasets and targets, non-temporal tree-based models, particularly CatBoost, consistently outperformed the temporal DL models. These findings suggest state-of-the-art temporal DL models still struggle with factors such as class imbalance, sparsity, irregularity, and asynchronicity of laboratory values that are typical of routinely collected laboratory data beyond the ICU, and that further research is needed to overcome these challenges.

## 1 Introduction

Electronic health records (EHRs) have become one of the primary sources of real-world data for machine learning (ML) applications in healthcare [1–5]. Clinical data, especially quantitative laboratory measurements, are collected in standardized clinical protocols and their numerical format provides the basis for computational and ML frameworks. These measurements are collected routinely in clinical practice and their longitudinal nature offers rich temporal information which can be effectively leveraged by temporal deep learning (DL) models to derive meaningful temporal patterns and improve predictive performance. Most existing temporal DL models applied to EHRs are designed for regularly sampled time-series [6–9], however, real-world clinical data are inherently inconsistent across patients with irregular, sparse, and incomplete observations. To address this limitation, various DL models have been proposed to handle irregular and multivariate time-series and successfully applied to longitudinal laboratory measurements in EHRs [10–13].

Independently of model architecture, most state-of-the-art temporal DL models have been developed and benchmarked primarily using established ICU EHR datasets and on relatively well-defined and balanced prediction targets such as mortality, readmission, and length of stay. While these are relatively common tasks for ICU admissions, they are less representative of prediction needs in general hospital settings [7, 10, 14–22]. Moreover, ICU data are characterized by relatively dense and frequent measurements, partly due to the higher monitoring intensity in critical care. In contrast, data from general hospital admissions are typically less frequent and sparser, as measurements are collected based on clinical needs rather than continuous monitoring. Given the predominance of ICU-focused benchmarks, there is thus only limited evidence on how temporal DL models perform in irregular and sparse non-ICU data settings, especially when applied to real-world highly imbalanced prediction targets.

To fill this knowledge gap, we selected the clinically highly relevant and yet under-explored problem in precision oncology to predict chemotherapy-related complications. Only a limited number of studies have addressed this problem through the lens of ML, and used only classical ML and no temporal DL models [23–26]. Specifically, we focus on predicting neutropenic fever and aplasia as two complications of chemotherapy, using routinely collected laboratory data from general hospital admissions. Aplasia and neutropenic fever are associated with chemotherapy-induced bone marrow suppression and both are considered serious and potentially life-threatening conditions. Early identification of patients at higher risk is essential to enable timely therapeutic interventions and improve outcomes [27–29].

To this end, we developed a training and evaluation pipeline including 7 state-of-the-art temporal DL models and 6 non-temporal classic ML models to predict chemotherapy-related complications from longitudinal laboratory measurements. The models were evaluated on an adult cohort extracted from the publicly available MIMIC-IV dataset [30], where neutropenic fever and aplasia occurred in 5% and 22% of patients, respectively, and an in-house pediatric cohort from the Department of Pediatrics and Adolescent Medicine of the University Hospital Erlangen (UKEr), with comparable disease incidence. These cohorts allowed us to evaluate the effectiveness of the temporal DL models under real-world clinical settings such as relatively small sample sizes, irregularly sampled data, and severe class imbalance, and to determine whether they provide predictive improvements over classical ML methods in challenging data scenarios.

## 2 Results

### 2.1 Study overview

Figure 1 provides an overview of our unified evaluation pipeline to compare classical ML models and temporal DL for clinical prediction tasks (details in “Methods”). The pipeline comprises three components: data extraction from EHR datasets, feature engineering of laboratory measurements into model-specific inputs, and standardized model training and evaluation. The data extraction module interfaces with the MIMIC-IV and UKEr datasets to retrieve the clinical cohorts and corresponding predictive targets. Laboratory measurements recorded within a predefined period before discharge are extracted as irregular, sparsely sampled time series (TS). The UKEr dataset contains 48 laboratory measurements, all of which were used for our study; for MIMIC-IV, we extracted the 100 most frequently measured items. Timestamps are available at minute-level resolution in MIMIC-IV and at daily resolution in UKEr. The feature engineering module then transforms the extracted TS of laboratory measurements into three alternative input formats, as required by the tested models: (i) regularly sampled, discretized, and imputed TS with indicators of missingness and time since last observation; (ii) irregularly sampled series with missingness masks and temporal gaps; and (iii) sequences of events represented as time–variable–value triplets.

**Fig. 1:**
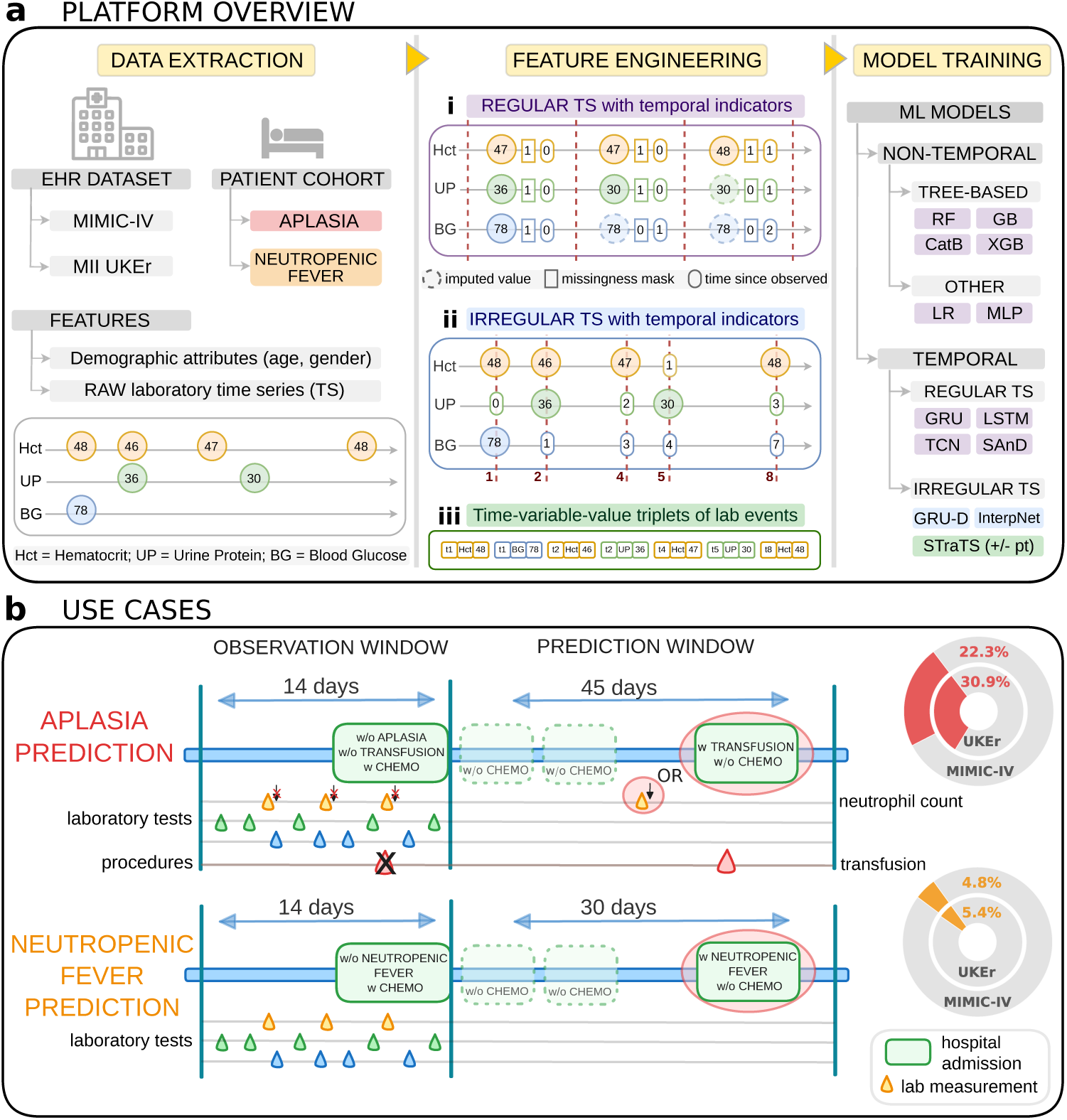
Methodology overview. (a) Pipeline for the systematic comparison of ML models. The data extraction module retrieves aplasia and neutropenic fever cohorts from the MIMIC-IV and UKEr datasets, extracting laboratory measurements as irregular and sparse TS. The feature engineering module produces model-specific inputs: (i) regularly sampled, discretized, and imputed TS; (ii) irregularly sampled series with similar indicators; or (iii) event triplets (time-variable-value). The model training module trains and evaluates all models. (b) Aplasia and neutropenic fever cohort extraction. Aplasia was defined by a transfusion procedure or low absolute neutrophil count, and neutropenic fever by concurrent diagnoses of neutropenia and fever. The target was to predict admission-level occurrence of the respective condition within the prediction window after discharge and before the next chemotherapy application, using data from the observation window starting 14 days before discharge.

The training module covers classical non-temporal ML models, temporal DL models designed for regular TS, and temporal DL models designed for irregular TS. The classical non-temporal ML models serve as a baseline to assess the benefits of DL architectures designed for temporal data. We included the following models: logistic regression (LR), multilayer perceptron (MLP), random forest (RF) [31], gradient boosting (GB) [32], extreme gradient boosting (XGB) [33], and CatBoost (CatB) [34]. All of them use input of type (i) (regular TS). As temporal DL models for regular TS, we included three general-purpose models — a gated recurrent unit (GRU) model [35], a long short-term memory (LSTM) model [36], and a temporal convolutional network (TCN) model [37] — as well as the SAnD model [17], which is specifically designed for clinical TS data. Also these models use input of type (i). Finally, we included three DL models designed for irregular TS data, all of which were specifically developed for longitudinal data extracted from EHRs: GRU-D [10], InterpNet [38], and STraTS [13]. GRU-D and InterpNet use input of type (ii) (irregular TS with temporal indicators), while STraTS is a transformer-based model that uses input of type (iii) (event triplets). All models were trained and evaluated via (nested) cross-validation (CV) schemes with hyperparameter tuning, and significance of differences in performance metrics computed on the test folds was assessed via statistical testing.

Cohorts comprised admissions of patients with a cancer diagnosis and at least one chemotherapy procedure, excluding cases in which the target could not be determined. Aplasia was defined by transfusion procedures or low absolute neutrophil count, and neutropenic fever by concurrent diagnoses of neutropenia and fever. The prediction target was the onset of the respective condition within the prediction window (45 days after discharge for aplasia, 30 days for neutropenic fever) and prior to the next chemotherapy cycle, while the observation window was set to 14 days prior to discharge. Applying these criteria yielded 4539 admissions in MIMIC-IV and 2446 in UKEr for aplasia (target observed in 22.25% and 30.94% of all admissions, respectively) and 5308 admissions in MIMIC-IV and 5210 in UKEr for neutropenic fever (target observed in 4.82% and 5.35% of all admissions, respectively).

### 2.2 Tree-based models outperform temporal deep learning models in predicting chemotherapy side effects

Table 1 shows test performances of the ML models quantified via the area under the receiver operating characteristic (AUC-ROC), the area under the precision-recall curve (AUC-PRC), and the F1 score (harmonic mean of precision and recall), obtained through a large-scale evaluation via a nested 5 × 3 CV scheme. The table summarizes results for 5 hyperparameter configurations per model, cohort, and target (one for each of the 5 outer splits in the 5 × 3 CV scheme), which were determined in the inner loop of the 5 × 3 CV that further splits the training fold of the corresponding outer loop into training and validation sets. Since splitting was carried out at the level of patients rather than admissions, this setup ensures that data from patients in the *i*^th^ test fold were used neither for hyperparameter selection nor for weight fitting of the models whose performances were evaluated on the *i*^th^ test fold, which is important to avoid data leakage in the evaluation scheme [39]. Effects of further design choices (temporal resolution of discretized TS, usage of temporal features in discretized TS, pretraining strategies for STraTS) were investigated in separate ablation studies whose results are reported in the following subsections.

**Table 1:**
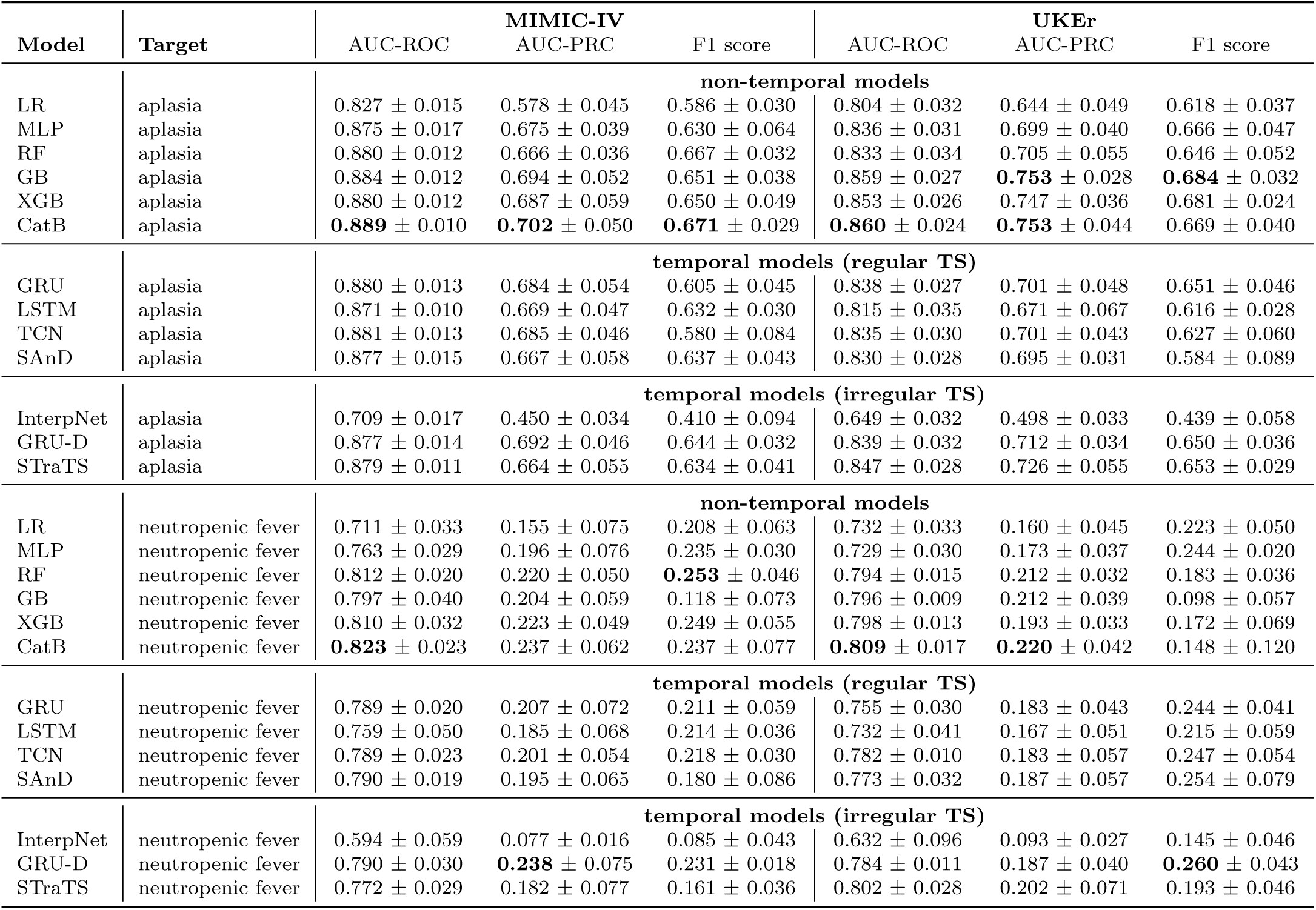
Performance of ML models on the aplasia and neutropenic fever cohorts from MIMIC-IV and UKEr datasets. The best score for each metric and cohort is highlighted in bold. Reported values correspond to means and standard deviations of test performances in the outer loop of a nested 5 × 3 CV scheme. Non-temporal models and temporal models for regular TS were trained on discretized data with daily resolution.

Overall, the occurrence of chemotherapy side effects was predicted with substantially above-random performance across all cohorts, reaching an AUC-ROC of 0.889 for aplasia and an AUC-ROC of 0.823 for neutropenic fever. Performance was higher for aplasia, likely reflecting the lower class imbalance for this outcome. CatBoost consistently emerged as the best-performing model across conditions and databases, achieving the highest AUC-ROC and top scores for at least two of three metrics in each dataset. Trained on a lossy daily-discretized representation, CatBoost still out-performed all temporal DL models for both regular and irregular TS, suggesting that modeling fine-grained irregular temporal patterns offers limited additional predictive value in this setting. InterpNet performed poorly across all metrics, likely because its interpolation-based approach struggles with the extreme sparsity of the data. LSTM models similarly underperformed, as their ability to capture long-range dependencies was limited by the short 14-day observation period. LR also showed lower performance, potentially due to the high dimensionality of the feature space resulting from concatenating values, missingness masks, and delta-time features. Excluding these three models, the remaining models showed a relatively narrow performance range, with AUC-ROC ∈ [0.875, 0.889] for aplasia and AUC-ROC ∈ [0.763, 0.823] for neutropenic fever on the MIMIC-IV dataset, and AUC-ROC ∈ [0.833, 0.860] for aplasia, and AUC-ROC ∈ [0.729, 0.809] for neutropenic fever on the UKEr dataset. Overall, these results suggest that most predictive information is preserved in the lossy, daily-discretized representation, and that this information is most effectively exploited by the non-temporal model CatBoost.

To assess the statistical significance of differences in model performance, pairwise tests were performed on metrics computed from five repetitions of 5-fold CV. Unlike for the computation of the results summarized in Table 1, we here used one fold-independent hyperparameter configuration per model, determined based on the mean validation performance in the inner loops of the nested 5×3 CV described above. While this setup has the disadvantage that validation and test data are not independent, it allowed us to obtain a sufficiently large number of performance scores per model, target, and cohort to enable statistical testing within the limits of our computational resources. Wilcoxon signed-rank tests with Benjamini–Hochberg correction show that CatBoost significantly outperforms all other models for both AUC-ROC (Figure 2a) and AUC-PRC (Figure 2b). The other three tree-based models also outperform most temporal models in these metrics, with only GRU-D achieving comparable AUC-PRC performance. Based on these two metrics, a clear ranking emerges: CatBoost ranks first, followed by the other tree-based models, then most temporal models (excluding LSTM and InterpNet, but on par with MLP), and finally LSTM, LR, and InterpNet. For the F1 score (Figure 2c), fewer significant differences are observed, consistent with the higher standard deviations reported in Table 1. Here, MLP, RF, and GRU-D achieve the highest scores, outperforming other models, followed by a heterogeneous group of temporal and non-temporal models (TCN, SAnD, GRU, XGB, CatBoost) with comparable performance. The divergence from AUC metrics likely reflects the F1 score’s sensitivity to classification thresholds and the precision–recall trade-off, whereas AUC-ROC and AUC-PRC are threshold-independent and capture the overall ranking of predicted probabilities.

**Fig. 2:**
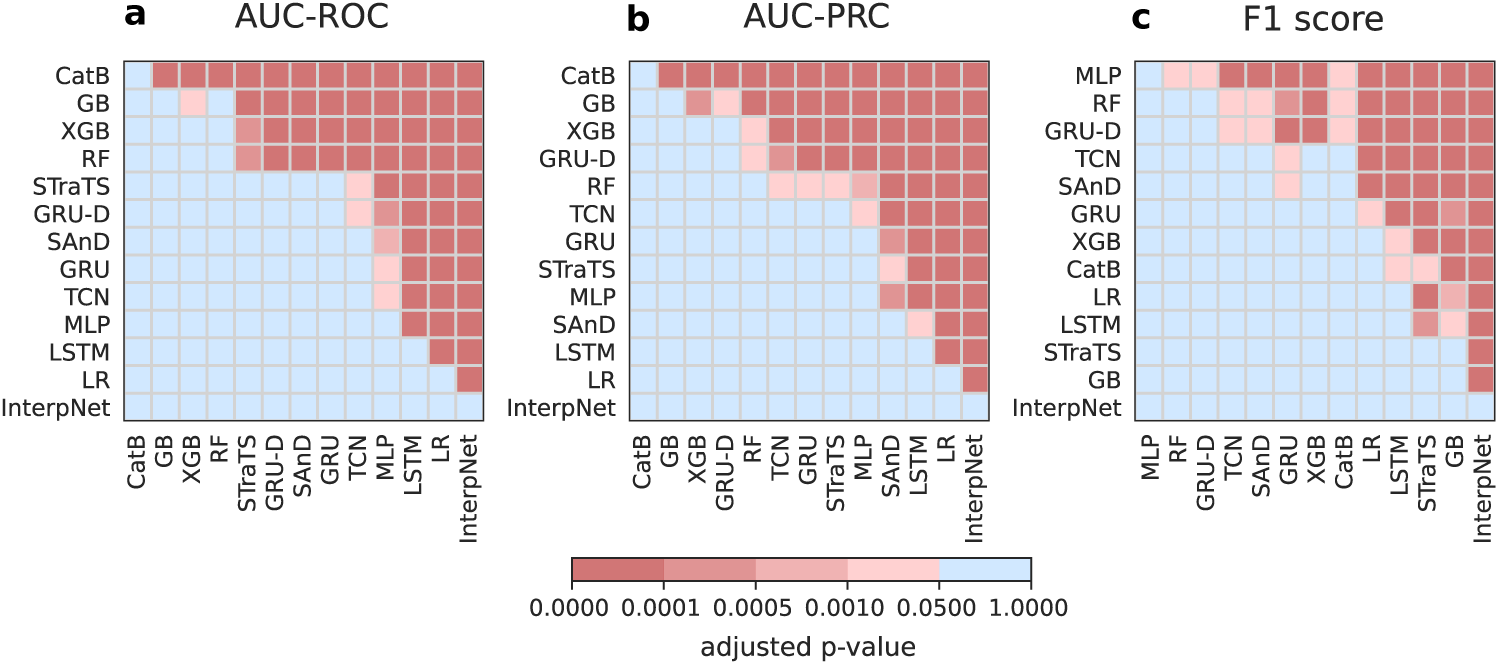
Statistical analysis of model performance across two cohorts from two EHR databases, with models evaluated using 5 runs of 5-fold CVs (4 datasets, 25 folds per dataset). *p*-values to assess statistical significance of differences in AUC-ROC (a), AU-PRC (b), and F1 scores (c) were computed with one-sided Wilcoxon signed-rank test and adjusted for multiple testing via Benjamini-Hochberg correction. In the heatmaps, models are ordered by their average performance for the corresponding metric.

### 2.3 Temporal features boost performance of non-temporal models

The use of three feature sets representing discretized and imputed values (V), missingness indicators (M), and temporal gaps (D) has become standard practice for training temporal DL models on regularly sampled sequences [10, 13]. However, non-temporal models are typically trained on V alone, without additional temporal indicators. To evaluate the contribution of temporal information in non-temporal models, these models were trained on all combinations of V, M, and D using nested 5 × 3 CV.

Test performances are reported in Figure 3. As expected, V is the most predictive single feature set, followed by D and M. This ordering reflects the fact that missingness can be inferred from the temporal gap, since a value of zero in D corresponds to an observed value in M, and vice versa. Notably, both M and D retain substantial predictive power, with average AUC-ROC values (across all models and cohorts) of 0.752 for M and 0.775 for D, compared to 0.814 for V, suggesting that the timing of measurement conveys important clinical information beyond the measured value.

**Fig. 3:**
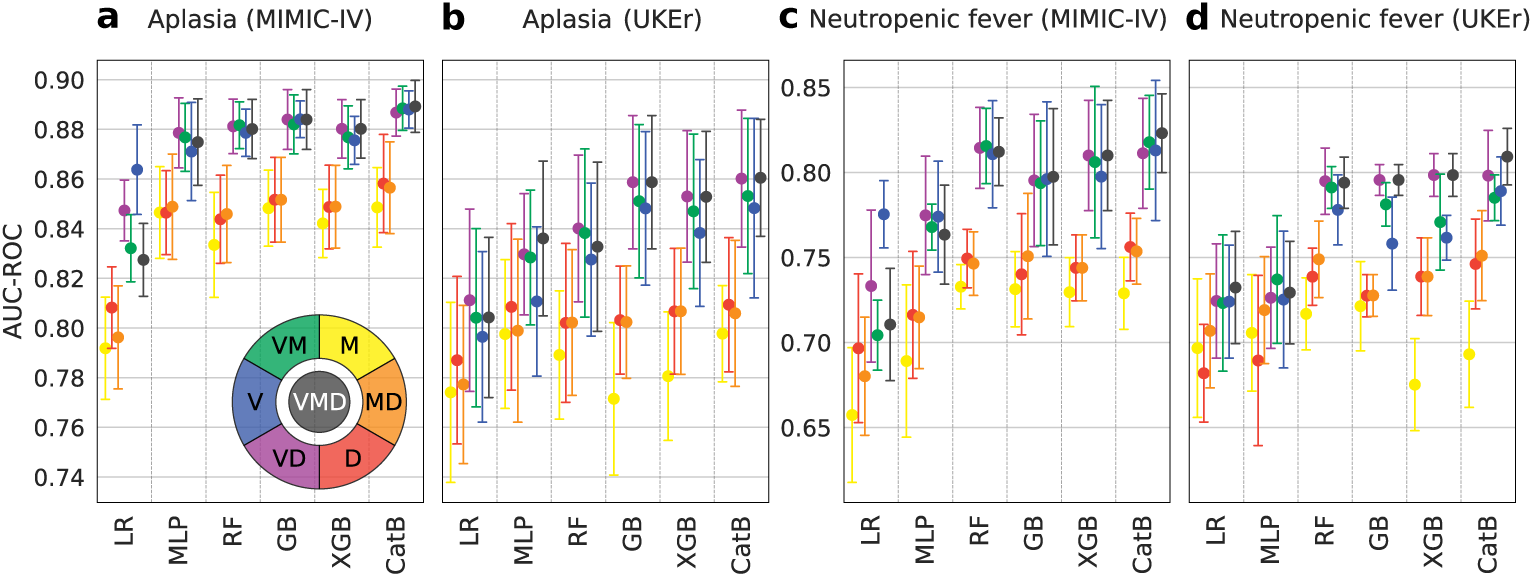
Performance of non-temporal models trained on all combinations of V (discretized imputed values), M (missingness indicators), and D features (temporal gaps since last observed) on four datasets: the aplasia cohort from MIMIC-IV (a) and UKEr (b), and the neutropenic fever cohort from MIMIC-IV (c) and UKEr (d). Plots show mean ± standard deviation of test performance across the outer folds of a nested 5×3 cross-validation scheme. Corresponding plots for AUC-PRC and the F1 score are shown in Figure S1.

All non-temporal models except LR benefit from the added temporal features. In CatBoost, the full V/M/D combination achieves the highest performance across all cohorts, likely due to its ability to leverage the categorical nature of M features. For RF, GB, and XGB, the best combination is V/D, possibly because these models can derive missingness information from D but lack native support for categorical features. In these models, V/M/D remains the most frequent second-best feature configuration and is never substantially worse than the best one.

### 2.4 Fine-grained temporal resolution does not improve predictive accuracy in sparse temporal data

Recent temporal modeling approaches emphasize capturing the precise timing of measurement events and handling irregular sampling intervals. In certain clinical settings, however, exact timestamps are either unreliable or even unnecessary, for instance if the physiological value remains stable over extended periods. In such cases, modeling exact timestamps can introduce noise or bias rather than improve predictive performance. This pattern is reflected in the empirical findings reported in Table 1. In the MIMIC-IV cohorts, the best-performing models are tree-based architectures trained on daily aggregated measurements, outperforming temporal models trained on exact timestamps. Performance in the UKEr cohorts, where timestamps were omitted due to their limited reliability, is comparable to that in the MIMIC-IV cohorts.

To systematically assess how temporal resolution influences predictive accuracy, non-temporal models and temporal DL models for regular TS were trained on data discretized at 3, 6, 12, and 24-hour intervals using nested 5 × 3 CV, with results presented in Figure 4. A 24-hour aggregation window yielded the best overall performance across both model classes, achieving the highest score in ten cases, followed by 12-hour (six cases) and 6- and 3-hour intervals (two cases each). Moreover, we trained the GRU-D and STraTS models designed for irregular TS with original timestamps retained or dropped, i.e., with all values for a given day set to the same time (Interp-Net was excluded from this comparison, as its interpolation-based mechanism depends on timestamps and daily alignment leaves too few points for interpolation). For both GRU-D and STraTS, removing timestamps did not significantly decrease performance, and the configuration without timestamps even performed better for one of the two targets.

**Fig. 4:**
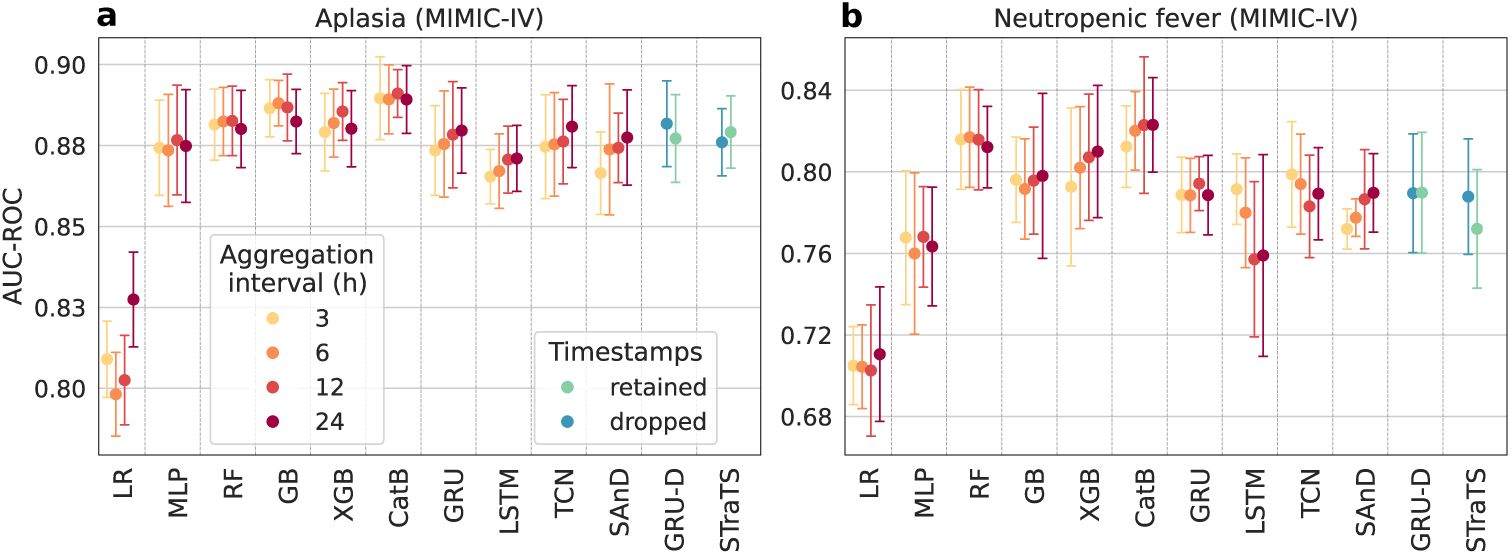
Performance of temporal models under varying temporal granularity in aplasia (a) and neutropenic fever (b) cohorts extracted from MIMIC-IV. Regular time-series models (GRU, LSTM, TCN, SAnD) were trained on data discretized at 3, 6, 12, and 24-hour intervals. Plots show mean ± standard deviation of test performance computed on the outer folds of a nested 5×3 cross-validation scheme. Irregular TS models (GRU-D and STraTS) were trained on data with original timestamps retained or with timestamps dropped (all values for a given day set to the same time).

Overall, these results indicate that, in the MIMIC-IV cohorts, daily aggregates provide predictive information equivalent to timestamped measurements. This suggests that longitudinal laboratory data without exact times such as our UKEr dataset provides sufficient temporal resolution for predicting aplasia and neutropenic fever. We hypothesize that this finding that daily trends are more informative than short-term fluctuations generalizes to other prediction targets that are expected to evolve over longer periods.

### 2.5 Pretraining does not consistently improve predictions of the temporal transformer model STraTS

Transformer-based DL architectures typically require large datasets to train effectively. When data are limited, pretraining on other data or a simpler task, followed by fine-tuning on the target cohort, can improve performance. To evaluate this approach, STraTS — the only transformer-based model included in our benchmark — was pre-trained using its native forecasting task either on the target cohort or on all admissions from the corresponding EHR database. Pretrained models were then fine-tuned, either fully or only on the fusion self-attention layer and forecasting head. As for statistical testing, we only used one hyperparameter configuration of STraTS for these experiments to keep resource requirements for pretraining and fine-tuning manageable.

For AUC-ROC (Figure 5a) and AUC-PRC (Figure 5b), none of the fine-tuning strategies consistently improved performance. Fine-tuning only the final layers generally reduced performance, particularly when pretrained on the target cohort, whereas full fine-tuning improved performance in only one dataset (MIMIC-IV neutropenic fever). Pretraining on the cohort with full fine-tuning, the approach used in the original STraTS study, yielded the best results among all configurations, although the improvements were minimal. For F1 scores (Figure 5c), performance improved across all cohorts and fine-tuning configurations, particularly when pretraining on the cohort and fine-tuning all layers, due to its sensitivity to the classification threshold. Fine-tuning made models more conservative in predicting positives, reducing false positives and increasing precision, while recall decreased slightly due to class imbalance, overall resulting in higher F1 scores. The limited benefit of pretraining is likely due to the size and sparsity of the considered cohorts. The forecasting pretraining task requires both a sufficient number of admissions and enough time points per feature, making the method less effective out of-the-box for our datasets, where only a few data points exist for many laboratory variables.

**Fig. 5:**
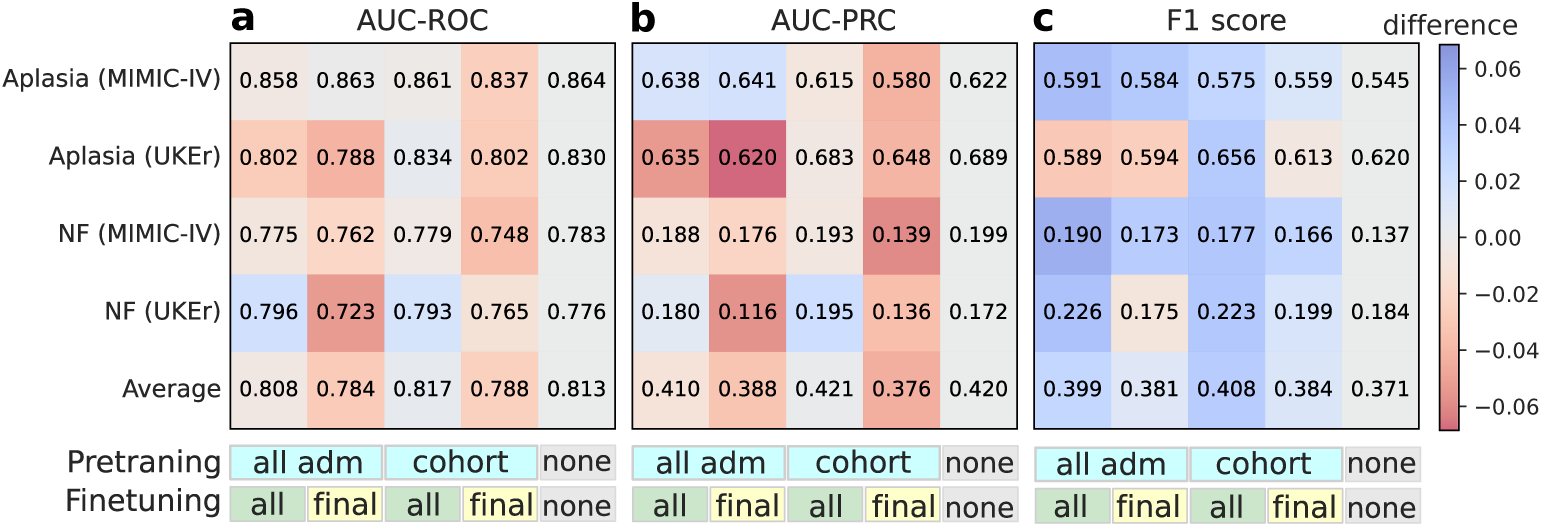
Performance of STraTS models, computed as mean AUC-ROC (a), AUC-PRC (b), and F1 score (c) from 5-fold CV, trained without pretraining (“none”) or under different pretraining configurations on aplasia and neutropenic fever (NF) cohorts. Models were pretrained by forecasting on the corresponding cohort (“cohort”) or on all suitable admissions from the corresponding database (“all adm”). Pretrained models were either fully fine-tuned (“all”) or fine-tuned only on the fusion self-attention layer and forecasting head (“final”). Heatmaps show the difference in performance between fine-tuned and non-pretrained models.

### 2.6 Interpreting key features in the best performing model

The reliability of the ML models predictions and their subsequent integration into real-world problems depend on the ability to move beyond the black-box predictions and explain whether the decisions are based on meaningful and domain-relevant information. To assess the clinical interpretability of CatBoost, our best performing model across all cohorts, we computed feature importances using CatBoost’s `PredictionValuesChange` metric, which measures the average absolute contribution of each feature to the model’s final predictions across all trees. Feature importances were computed for models trained on the full V/M/D features, allowing us to analyze the relative contribution of each feature set and their distribution across the 14-day observation period. The importance scores were averaged across the 5-fold CV.

Figure 6 shows the results for the neutropenic fever cohort from the UKEr dataset (results for the other three cohorts are similar and shown in Figures S2–S4). V features dominate the top predictors (Figure 6a, b), indicating that measured laboratory values contain the primary predictive information. In all four cohorts, the D feature set (temporal gaps) also contributed to the most important features (Figure 6b and Figures S3b, S4b, S5b), highlighting the importance of including these features to capture sparsity and informative missingness patterns. M features consistently yielded very low importance scores confirming that the missingness can be inferred from temporal gaps and these features did not add additional predictive value.

**Fig. 6:**
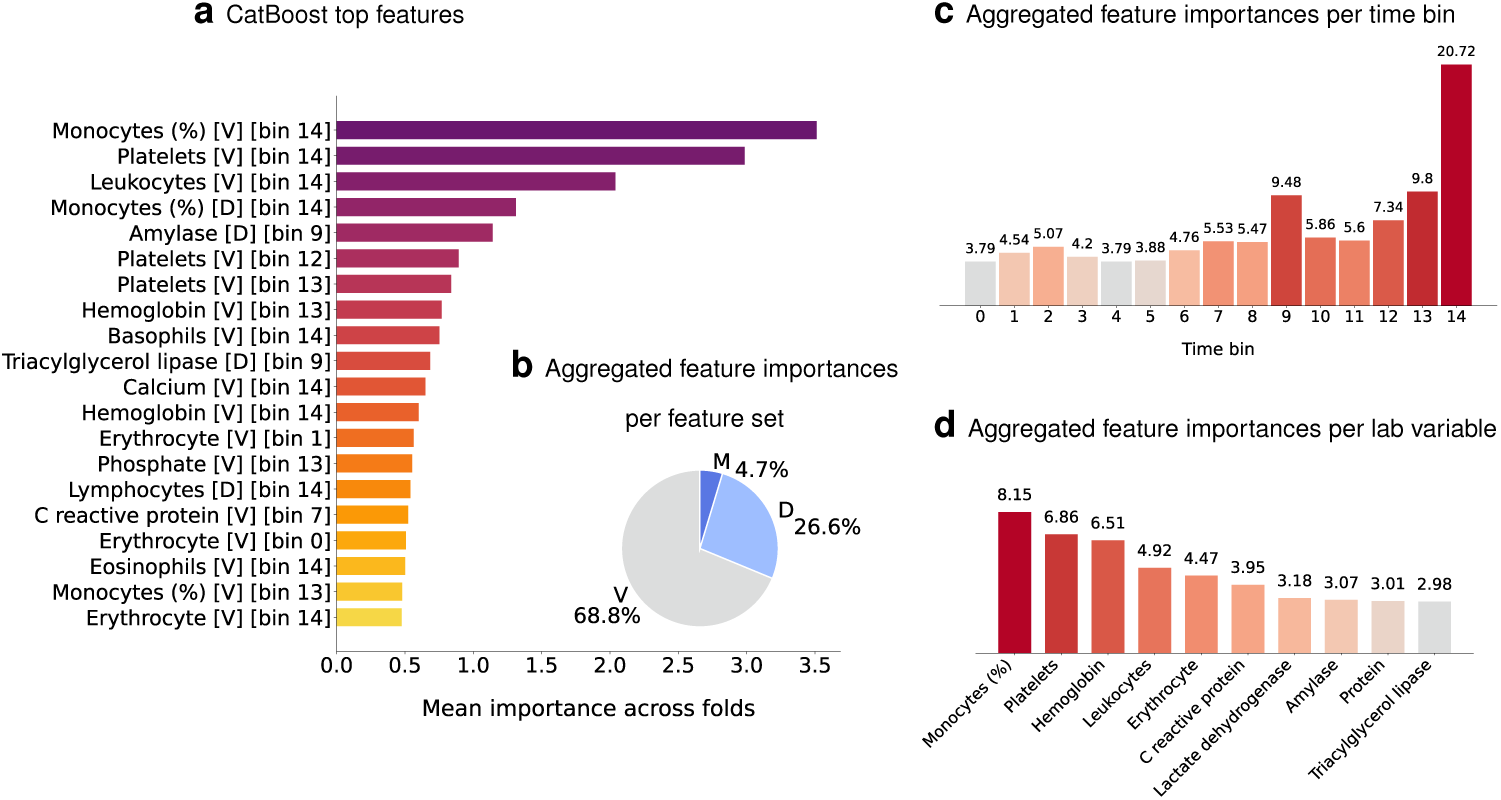
Analysis of CatBoost feature importance scores for the neutropenic fever cohort from the UKEr dataset (V/M/D features, daily resolution, 5-fold CV). (b–d) Contributions of feature sets (b), time bins (c), and laboratory variables (d) assessed by summing the importance scores for each feature type within its respective category.

As shown in Figure 6c, there is a consistent temporal pattern in time bin distribution. Features from the most recent time bins (closest to discharge) had a higher importance than earlier measurements. This suggests that measurements closer to discharge may already capture early changes associated with chemotherapy effects, which makes them more informative for predicting neutropenic fever. We observe similar patterns for the MIMIC-IV neutropenic fever cohort (Figure S3c) and the two aplasia cohorts (Figures S4c and S5c).

Analyzing the effect of the laboratory variables across V, M, and D representations and time bins (Figure 6d and Figures S3d, S4d, S5d) revealed that hematologic measurements were the most influential predictors of both neutropenic fever and aplasia. White blood cell-related features such as monocytes, neutrophils, and leukocytes are the early indicators of bone marrow suppression and a compromised immune system. In the UKEr cohorts (Figure 6d and Figure S2), red blood cell–related features such as hemoglobin and erythrocyte counts, along with platelet measures, had higher importance scores, suggesting a broader underlying bone marrow dysfunction.

Non-hematologic features such as electrolytes, metabolic markers, and enzymatic parameters also contributed but generally ranked lower suggesting that the model primarily relied on hematologic indicator. In MIMIC-IV cohorts (Figures S3 and S4), uric acid was one of the highest ranked features, suggesting additional biological mechanisms that need to be explored further. Taken together, these findings demonstrate that the models make their predictions based on plausible and clinically meaningful signals. While the feature importance confirms the expected hematologic predictors, the prominence of metabolic related markers opens avenues for exploring additional affected pathways.

## 3 Discussion

The release of large intensive care EHR databases such as MIMIC-III [40], along with other curated ICU datasets provided through competitions like the PhysioNet Challenges 2012 and 2019 [41, 42], has driven the development of numerous DL models for laboratory TS classification, most of which focus on handling the irregular and multivariate nature of laboratory data [10, 13, 38, 43]. However, these models were largely developed and tested on these established ICU EHR datasets, using a narrow set of ICU-specific classification tasks including in-hospital mortality, readmission, and length of stay. Moreover, baselines in these studies are often limited to other DL architectures, while tree-based methods have largely been phased out, as they did not outperform DL models on these specific ICU-specific tasks.

With the release of MIMIC-IV — which includes a hospital module covering over 500K general admissions alongside 95K ICU admissions — general-admission data has become more widely available. Unlike ICU data, general-admission data does not include vital signs measured continuously or frequently by bedside monitors or sensors, and typically reports sparser laboratory measurements. Nevertheless, research has largely remained focused on the same ICU benchmarks from MIMIC-III and PhysioNet, likely to enable direct comparison with prior work and to leverage established preprocessing practices for these datasets, which have not yet been consolidated for MIMIC-IV [44–50]. Self-supervised modeling efforts aimed at deriving admission-level representations of laboratory data for multiple downstream tasks have also largely relied on ICU datasets [47, 51, 52]: the only self-supervised model trained on general admission data from MIMIC-IV we are aware of is designed for imputation rather than predictive modeling [53]. Consequently, the performance of existing predictive models on longitudinal EHR data from general admissions remained largely unexplored.

In this study, we closed this knowledge gap by training and comparing a comprehensive set of ML models on cohorts extracted from general admissions of two EHR databases, focusing on two chemotherapy side effects as prediction tasks. Surprisingly, tree-based models consistently outperformed temporal DL models, especially when given access to temporal M/D features that allow them to capture sparsity patterns and informative missingness. In contrast to the picture emerging from the ICU-centric literature, this shows that temporal DL models are not universally superior to simpler models and underscores that model performance is context-dependent.

The main limitation of our study is that it remains unclear *why* the tree-based models outperformed the temporal DL models. Drawing on an analogy to recent bench-marking studies for tabular data which have shown that tree-based models are often better equipped to handle various dataset irregularities than DL models [54, 55], we hypothesize that several factors may contribute:

- Weak temporal signal: In EHR data from general admissions, the temporal signal can be weak or limited to a subset of features, with many spurious fluctuations. Laboratory values often vary slowly, so models trained on daily patterns without sequential awareness perform adequately, and simple forward/backward imputation can approximate missing values effectively. Feature sparsity further complicates modeling, with some variables measured only once per admission, while others are recorded multiple times with clinically relevant variation. In this scenario, treating all features as temporal can dilute the overall predictive signal.
- Temporal granularity: Efforts to model irregular TS often overlook that general-admission timestamps can be noisy, for instance, when they reflect the time samples were processed rather than collected. For slowly changing variables, precise timing is often less relevant, as measured values remain valid over extended intervals. Tree-based models can implicitly accommodate different resolutions by using data from a single or multiple time points, whereas transformers for TS data such as STraTS focus on modeling exact timestamps.
- Informative missingness: Patterns of missing data not at random can carry predictive signals that reflect domain constraints (e. g., labs analyzed in panels), value thresholds (e. g., labs omitted if another measurement falls below a detection limit), or decisions of clinicians (e. g., labs ordered based on patient assessment). Tree-based models can naturally exploit these signals, as they natively handle binary variables and can split directly on the missingness features (particularly CatBoost). In contrast, temporal neural networks rely on hidden-state propagation, which may dilute or forget sparse missingness signals.
- Value resolution: Laboratory measurements are typically interpreted relative to normal ranges, and small variations within these ranges are often clinically insignificant. Tree-based models naturally handle this by splitting data at thresholds, effectively capturing clinically relevant intervals, whereas DL models evaluate individual values.

These observations suggest multiple avenues for improving temporal DL for general-admission EHR data: Firstly, temporal DL models could be improved by gauging the temporal dynamics of each feature, e. g., by defaulting to non-temporal modules for very sparse sequences or by accommodating varying degrees of temporality across features. Recent pretraining methods have begun to address this issue by proposing time-sensitive contrastive learning and masking to capture varying data density [51], but they have not been evaluated at the sparsity levels typical of general-admission data. Secondly, learning the appropriate temporal resolution for each feature and assigning measured values to the corresponding interval can help to reduce noise arising from modeling exact timestamps. Recent efforts to produce multi-scale representations have employed warping modules that adaptively unify irregular TS at a given scale, and have stacked multiple warping and attention modules to learn across different scales [56]. However, by learning shared temporal scales across all features, these methods may not yet detect the most appropriate temporal resolution for each variable. Thirdly, it may be possible to improve temporal DL models by separately modeling value and missingness features or by employing temporal attention to explicitly capture missing events. Recently, pretraining strategies have been proposed that incorporate missingness into attention mechanisms [47], but they still do not natively handle irregular TS. Finally, we hypothesize that incorporating reference ranges from the literature or learning population-specific intervals to reduce the impact of within-range variation could improve the performance of temporal DL models.

Overall, our study highlights the potential of general-admission EHR data, a relatively underexploited resource, to address complex clinical questions. At the same time, it exposes limitations in current temporal DL models, stemming from the assumptions they make and the limited and strongly ICU-focused evaluation they underwent in previous studies. To advance the development of predictive models for longitudinal EHR data that perform well under sparse temporal conditions that align with clinical practice where measurements are often reduced to minimize costs and patient discomfort, we thus suggest two concrete measures: Firstly, new models should be tested on EHR data from general admission cohorts such as MIMIC-IV’s general admission module, using prediction tasks that are relevant also beyond the ICU. Secondly, new models should always be tested against well-tuned tree-based baselines trained on combined sets of V/M/D features.

## 4 Methods

### 4.1 Electronic health record databases

We used two independent EHR databases: MIMIC-IV and the pediatric subset from the Oncology Module of the Core Dataset provided by the UKEr node within the Medical Informatics Initiative Germany (MII) infrastructure. MIMIC-IV is a publicly available EHR database containing de-identified information on patients admitted to the Beth Israel Deaconess Medical Center between 2008 and 2019 [30]. It includes demographics, diagnoses, procedures, laboratory tests, and medications, and is organized into two main modules: HOSP, sourced from the hospital-wide EHR, and ICU, sourced from the in-ICU clinical information system. For our study, we extracted admissions from the HOSP module to evaluate laboratory-based prediction tasks across general hospital admissions. This module includes 223452 unique patients and 546028 hospitalizations. The dataset from the MII node at UKEr provides pseudonymized patient information, including demographics, diagnoses, procedures, laboratory tests, and hospital admissions, collected at UKEr’s Department of Pediatrics and Adolescent Medicine between 2012 and 2024 as part of the MII. It comprises 37586 patients and 79096 admissions. Note that patient demographics differ between the two datasets (Figure 7): While the MIMIC-IV’s HOSP module mainly contains data from adult patients, the UKEr dataset exclusively contains data from pediatric patients.

**Fig. 7:**
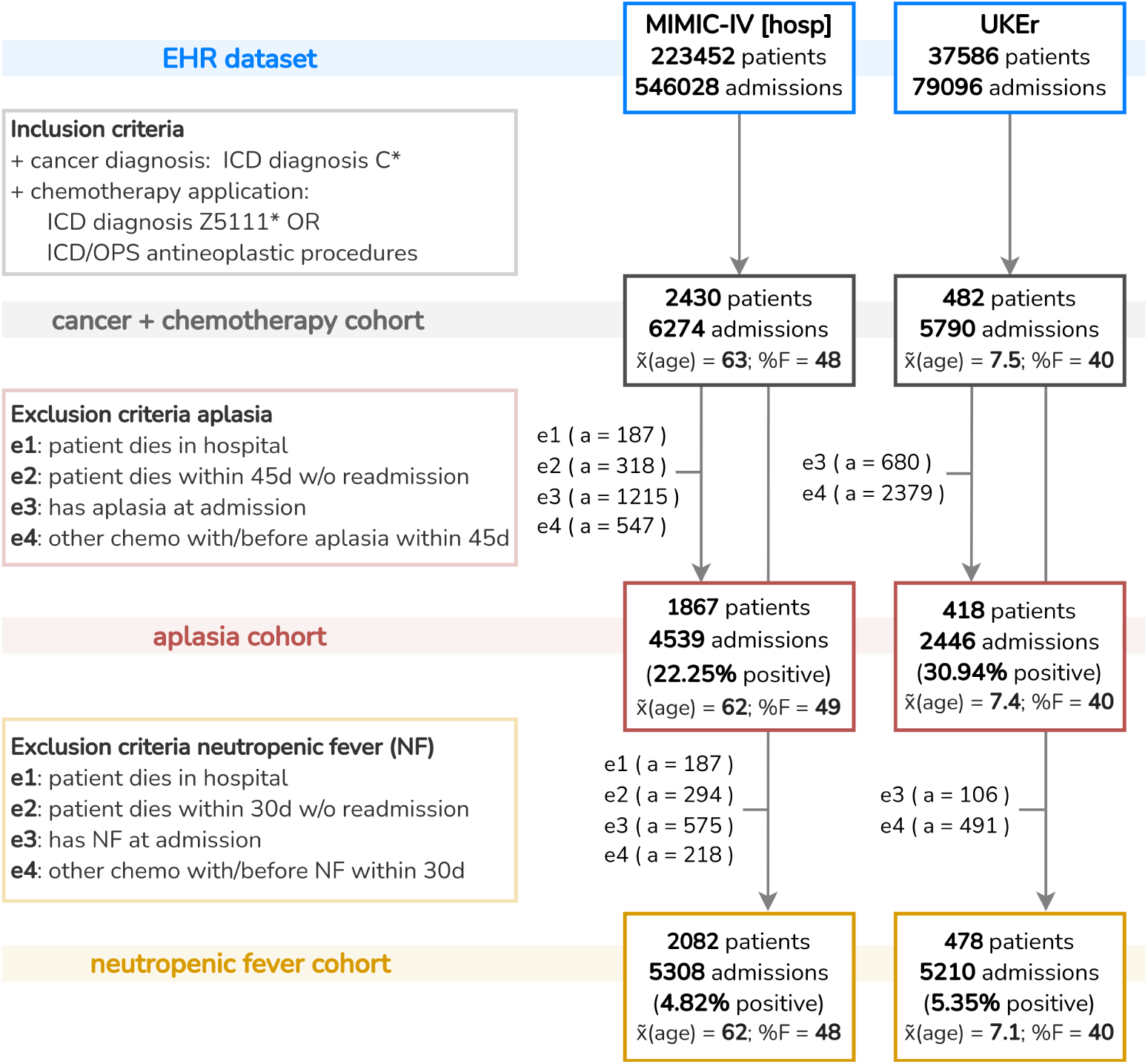
Flowchart of the cohort extraction procedure for aplasia and neutropenic fever in MIMIC-IV and UKEr, showing inclusion and exclusion criteria and the number of admissions retained. For each extracted cohort, the number of admissions, the number of patients, the median age *x̃*(age), and the fraction of female patients (%F) are reported. Exclusion criteria e1 and e2 were not applied in UKEr as death information was not available. Note: a single admission may meet more than one exclusion criterion.

### 4.2 Cohort extraction

Cohorts for aplasia and neutropenic fever were retrospectively extracted from the MIMIC-IV and UKEr EHR databases. A cohort comprising cancer patients who had received chemotherapy (“cancer + chemotherapy cohort”) was extracted according to predefined inclusion criteria, and the two outcome-specific subcohorts were then derived by applying exclusion criteria. Figure 7 summarizes the extraction procedure and reports the number of patients and admissions retained at each step. Patients were included in the cancer + chemotherapy cohorts if they had a cancer diagnosis (ICD-10 codes beginning with “C”) and if there is evidence of chemotherapy administration based on the most specific available diagnosis and procedure codes. In UKEr, chemotherapy was defined by the ICD-10 diagnosis codes “Z511” (chemotherapy session for a neoplasm) or “Z512” (other chemotherapy), or operation and procedure (OPS) codes beginning with “8-542” (non-complex chemotherapy), “8-543” (moderately complex and intensive block chemotherapy), or “8-544” (highly complex and intensive block chemotherapy). In MIMIC-IV, chemotherapy was identified via the ICD-10 diagnosis code “Z5111” (encounter for antineoplastic chemotherapy) or via ICD-10 procedure codes corresponding to intravenous, arterial, subcutaneous, intrathecal, or epidural administration of antineoplastic agents (full list in Table S1). For the prediction tasks, the observation window was set to 14 days before discharge. Prediction windows were set to 30 days after discharge for neutropenic fever and 45 days for aplasia. Aplasia was defined as either an absolute neutrophil count below 500 µL (itemid “52075” in MIMIC-IV, LOINC code “751-8” in UKEr) or by transfusion of red blood cells or platelets (identified by ICD procedure codes in MIMIC-IV and OPS codes in UKEr, which are listed in Table S1). Neutropenic fever was defined as the concurrent presence of ICD codes for neutropenia (codes starting with “D70”) and fever (codes starting with “R50”) within the same admission. Outcome-specific subcohorts of the cancer + chemotherapy cohorts were derived by excluding admissions for which the target could not be clearly determined. Exclusion criteria were:

- Exclusion criterion e1: Patient died in hospital (no discharge, hence no prediction possible).
- Exclusion criterion e2: Patient died within the prediction period without readmission (insufficient information to determine whether the condition developed).
- Exclusion criterion e3: Patient had the condition during the current admission (unclear whether the condition was pre-existing or induced by the latest chemotherapy).
- Exclusion criterion e4: Patient underwent another chemotherapy during the prediction period and developed the condition during or after that admission (unclear whether the condition was due to the initial or subsequent chemotherapy).

All criteria were applied to MIMIC-IV, while only e3 and e4 were applied to UKEr due to unavailable mortality information. After applying the exclusion criteria, the final aplasia cohorts comprised 4539 admissions in MIMIC-IV and 2446 in UKEr. Aplasia occurred within the 45-day prediction window in 22.25% and 30.94% of admissions, respectively. For neutropenic fever, the cohort extraction process yielded 5308 admissions in MIMIC-IV and 5210 in UKEr. Neutropenic fever occurred within the 30-day prediction window in 4.82% and 5.35% of admissions, respectively.

### 4.3 Feature extraction

Demographic data (age and sex at birth) and laboratory measurements up to 14 days before discharge were collected for each admission. The UKEr dataset includes 48 LOINC laboratory items, all of which were included in the analysis. In MIMIC-IV, 660 distinct laboratory items occur in the cancer + chemotherapy cohort, but many of them in only very few admissions. Thus, the 100 most frequently recorded item IDs (present in at least 6% of admissions) were selected for analysis. MIMIC-IV laboratory measurements include precise timestamps, whereas UKEr records only dates, with most items measured at most once per day (for admissions with several measurements of the same item on the same day, we used daily averages).

### 4.4 Data representations

Laboratory measurements were represented using three input types, suitable for different models: (i) regularly sampled, discretized, and imputed TS with indicators of missingness and time since last observed, for non-temporal models and temporal models for regular TS; (ii) irregularly sampled series with missingness masks and temporal features for irregular TS models GRU-D and InterpNet; and (iii) sequences of events represented as time–variable–value triplets for the transformer-based model STraTs.

For input type (i), laboratory measurements were mean-aggregated over 3, 6, 12, and 24-hour intervals, and imputed using forward-fill followed by backward-fill. Any remaining missing values were zero-imputed. Alongside these imputed values (denoted as V), two sets of temporal features were computed to capture sparsity, following the approach suggested in [10]: a missingness mask (M) and time since last observation (D). Formally, for a laboratory item *d* in an admission, let *X^d^* = (*x*_1_*, …, x_t_, …, x_T_* ) denote its univariate TS over *T* discrete time bins, with *x_t_* representing the mean of all measured values for *d* during the time span represented by *t*. Note that *X^d^* may contain missing entries *x_t_* = NA. The derived feature sets *V ^d^* = (*v*_1_*, …, v_t_, …, v_T_* ), *M^d^* = (*m*_1_*, …, m_t_, …, m_T_* ), and *D^d^* = (*δ*_1_*, …, δ_t_, …, δ_T_* ) are defined as follows:

- Values (V): *v_t_* is defined as *v_t_* = *x_t_* if *x_t_* ≠ NA. If *x_t_* = NA, we defined *v_t_* as *v_t_* = *x_t__L_*, where *t_L_* is the latest time bin before *t* with *x_t__L_* = NA if such a time bin exists (forward fill), as *v_t_* = *x_t__R_*, where *t_R_* is the earliest time bin after *t* with *x_t__R_* ≠ NA if such a time bin exists but *t_L_* does not exist (backward fill); and as *v_t_* = 0 if *x_t_* = NA for all time bins *t*.
- Mask (M): *m_t_* is defined as *m_t_* = 1 if *x_t_* ≠ NA and as *m_t_* = 0 otherwise.
- Temporal gap (D): *δ_t_* is defined as *δ_t_* = *δ_t_^′^/T*, where *δ_t_^′^* is defined recursively as follows:

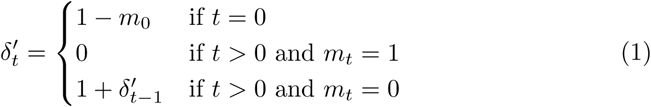

All possible combinations of these feature sets (V, M, D, V/D, V/M, M/D, V/M/D) were concatenated to form the input representation for the models. Note that our definition of the D features slightly differs from the original formulation [10] which always uses *δ*_0_ = 0. Our formulation distinguishes series with and without observations at the first bin and can thus be interpreted as a lower bound on the number of bins since the last observation.

For input type (ii), each laboratory TS is represented using three input features, similar to input (i), but now defined at exact timestamps rather than aggregated over fixed time intervals. Formally, for an admission, let *S* = (*s*_1_*, …, s_t_, …, s_n_*) denote the ordered time stamps (in minutes since the start of the observation window) for which measurements were observed across all laboratory tests for that admission, and let *s_max_* denote the maximum possible timestamp that can be recorded across all admissions (depending on the observation window). For a laboratory item *d*, let *X^d^* = (*x*_1_*, …, x_t_, … x_n_*) denote the recorded values at timestamps *S*, where some entries may be missing. The derived value features *V ^d^* = (*v*_1_*, …, v_t_, …, v_n_*) are defined as *v_t_* = *x_t_* if *x_t_* ≠ NA and *v_t_* = 0 otherwise (note, here the missing values are only zero-imputed). The missingness mask *M^d^* = (*m*_1_*, …, m_t_, …, m_n_*) is defined as in input (i), although the indices *t* now refer to exact timestamps rather than time windows. As the third feature set, InterpNet encodes temporal information by directly using the timestamps *S*, normalized by *s_max_*. GRU-D, on the other hand, uses temporal gaps *D^d^* = (*δ*_1_*, …, δ_t_, …, δ_n_*) since last observation, where *δ_t_* = *δ_t_^′^/s_max_* and *δ_t_^′^* is computed recursively as follows:

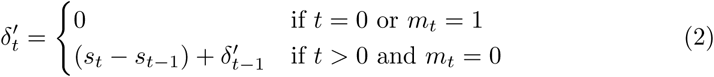

For input type (iii), each measurement *m_i_* in an admission is encoded as the tuple *m_i_* = (*s_i_, f_i_, x_i_*), where *s_i_* is the timestamp in minutes, *f_i_* is an integer index corresponding to the laboratory variable, and *x_i_* is the measured value.

Patient demographics were encoded as follows: For logistic regression and tree-based models, demographic features were directly concatenated with laboratory features prior to training. For all other models, demographic features were first transformed via a linear layer to create a demographic embedding, which was then concatenated with the temporal embedding derived from laboratory features to form the combined representation used for prediction.

### 4.5 Tested machine learning models

We tested 6 non-temporal ML models, 4 temporal DL models for regular TS, and 3 temporal models for irregular TS. The model characteristics are described in Table 2.

**Table 2:** Overview of tested models, classified by type of temporal handling (NT: non-temporal model, RTS: DL model for regular TS, ITS: DL model for irregular TS), model scope (whether developed for EHR data or not), and input data representation (among those visualized in Figure 1a and described in Section 4.4).

| Name | Abbr. | Short description | Type | EHR | Input |
| --- | --- | --- | --- | --- | --- |
| Linear regression | LR | Linear model for binary outcomes via a logistic function | NT | no | (i) |
| Multi-layer perceptron | MLP | Feedforward network with fully connected layers and nonlinear activations | NT | no | (i) |
| Random forest | RF | Ensemble of decision trees to reduce overfitting | NT | no | (i) |
| Gradient boosting | GB | Sequential weak decision trees correcting previous errors | NT | no | (i) |
| XGBoost | XGB | Optimized gradient boosting with regularization and parallelization | NT | no | (i) |
| CatBoost | CatB | Gradient boosting optimized for categorical features | NT | no | (i) |
| Gated recurrent unit | GRU | RNN with update and reset gates to mitigate vanishing gradients | RTS | no | (i) |
| Long short-term memory | LSTM | RNN with memory cells and gates to capture long-range dependencies | RTS | no | (i) |
| Temporal convolutional network | TCN | Network with dilated causal convolutions and residual connections | RTS | no | (i) |
| Self-attention network for diagnosis | SAnD | Masked self-attention with positional encoding for sequence segments | RTS | yes | (i) |
| GRU with decay | GRU-D | GRU variant with trainable decay to model irregular temporal gaps | ITS | yes | (ii) |
| Interpolation network | InterpNet | Semi-parametric model interpolating features to fixed reference points | ITS | yes | (ii) |
| Self-attention transformer for TS | STraTS | Transformer using attention to aggregate event triplets | ITS | yes | (iii) |

### 4.6 Cross-validation schemes used for model evaluation

The performance scores reported in Table 1 were obtained using V/M/D features with daily resolution for models with input type (i). They were determined via nested CV with a 5-fold outer loop to produce five 4:1 outer splits into training and test data and a 3-fold inner loop that further splits the training data from the outer fold into three partitions of training and validation data with size ratio 2:1. Both inner and outer splits were stratified by the target and grouped by patient to ensure all admissions from a single patient remained in the same fold.

For temporal models, the search grid was based on top-performing configurations from previous work [13], expanded around the reported best parameters. The optimal architecture per cohort and outer fold was selected based on the highest average AUC-ROC across all inner folds, followed by retraining of the best architecture on the entire training data from the outer split. For DL models, the number of training epochs was determined via early stopping monitored on the validation set of the inner split, using the mean over the three inner folds for retraining of the best performing architecture. For the ablation studies reported in Sections 2.3 and 2.4, we applied the same nested × 3 CV setup, training and testing models on feature sets that varied in aggregation interval or composition of V, M, and D features.

For the statistical analysis of differences in performance scores (Figure 2), we used only one hyperparameter configuration per model and cohort, which we selected based on mean AUC-ROC across all 5×3 = 15 inner folds of the nested CV scheme explained above (using V/M/D features with daily resolution for models with input type (i)). For DL models, we again set the number of training epochs to the mean number of epochs of the best configuration determined via early stopping on the inner folds. With this fixed configuration, we trained 25 models per cohort on the training sets of five repetitions of a 5-fold CV and then assessed model performances on the corresponding test sets.

To assess the effect of pretraining on STraTS (Section 2.5), we pretrained STraTS models via their inherent forecasting task on the last 14 days before discharge, again using the overall best performing hyperparameter configuration from the nested 5 × 3 CV. The maximum observation window was set to 7 days with a 24-hour forecasting horizon, adjusted for the sparser laboratory data in general hospital admissions. Pretraining was performed either on the training sets of a 5-fold CV of the corresponding cohort, or on all admissions in the EHR databases, excluding those for patients included in the target cohort. This allowed pretraining on the full EHR to be done only once per cohort, as repeating it for each fold would have been too computationally intensive. To determine the number of epochs for pretraining, we further split the training sets of the 5-fold CV into training and validation sets with size ratios 4:1, pretrained STraTS models on the smaller training sets with early stopping monitored on the validation set, and then again pretrained on the entire training sets from the 5-fold CV for the same number of epochs. Fine-tuning was performed either on all layers or only the last layers (fusion self-attention and forecasting head), following the same early stopping and retraining procedure.

### 4.7 Performance metrics

Predictive performance was assessed using the area under the receiver operating characteristic curve (AUC-ROC), the area under the precision-recall curve (AUC-PRC), and the F1 score, suitable for imbalanced tasks. Computational performance included memory usage and runtime during training and inference. For classical models, memory was measured as the process memory consumed by the fit and predict methods. For neural networks, peak memory was recorded per batch. Additional metrics for neural networks included training time per epoch and total number of parameters. Models were trained on an NVIDIA RTX A6000 (51 GB, CUDA 8.6). Tree-based models (except CatBoost) and LR ran on CPU, while all others used GPU acceleration. Memory usage was monitored using the Memory Profiler package (https://github.com/pythonprofilers/memory profiler) for CPU and PyTorch memory allocation functions for GPU. Execution time was measured using Python’s built-in time module. The resulting computational metrics across models and cohorts are reported in Figure S1.

### 4.8 Implementation details

All models were implemented and evaluated within a consistent computational framework. For classical ML models, we used established Python packages: RandomForestClassifier (RF), HistGradientBoostingClassifier (GB), and LogisticRegression (LR) from the scikit-learn package (https://scikit-learn.org/); CatBoostClassifier (CatB) from the catboost package (https://catboost.ai/); and XGBClassifier (XGB) from the xgboost package (https://xgboost.ai/). MLP, GRU, and LSTM models were implemented in PyTorch (https://pytorch.org/), while other neural network models (TCN, SAnD, GRU-D, InterpNet, and STraTS) were adapted from the STraTS pipeline (https://github.com/sindhura97/STraTS), which includes the pro-posed STraTS models as well as implementations of other baselines based on the original repositories: TCN (https://github.com/locuslab/TCN), SAnD (https://github.com/khirotaka/SAnD), InterpNet (https://github.com/mlds-lab/interp-net), and GRU-D (https://github.com/PeterChe1990/GRU-D). For these models, feature engineering, data loading, model training, and evaluation were reimplemented to ensure consistency with the procedures used for classical models. GPU-enabled training was conducted using PyTorch (CUDA 11.7). For classical models, hyperparameter search in the inner loop of nested cross-validation was conducted using GridSearchCV from scikit-learn. For neural network models, cross-validation setups were explicitly developed to allow flexibility, replicating the procedure for classical models when comparing models, or enabling dedicated CV setups, for instance in pretraining-finetuning scenarios. Statistical analysis was performed using Wilcoxon’s test from SciPy (https://scipy.org/), with multiple testing correction from statsmodels (https://www.statsmodels.org/).

## Availability of data and materials

- The MIMIC-IV dataset was accessed under a data use agreement through the PhysioNet project. Access to this dataset requires prior approval and completion of the relevant data use agreement and can be requested here: https://physionet.org/content/mimiciv/3.1/.
- The in-house UKEr dataset was analyzed in de-identified form on a server hosted within the network of the UKEr. The dataset is part of the Core Dataset of the Medical Informatics Initiative Germany (MII). Access can be requested through the German Portal for Medical Research Data (FDPG): https://forschen-fuer-gesundheit.de/en/.
- The source code underlying this study, including code for cohort extraction, data preprocessing, and model training, is available on GitLab and can be accessed via the following link: https://github.com/bionetslab/ChemoTreeVsDL.

## Data Availability

The MIMIC-IV dataset is publicly available and access to it requires prior approval and completion of the relevant data use agreement.
The University Hospital Erlangen (UKEr) dataset is part of the Core Dataset of the Medical Informatics Initiative Germany (MII). Access can be requested through the German Portal for Medical Research Data (FDPG).

https://physionet.org/content/mimiciv/3.1/

https://forschen-fuer-gesundheit.de/en/

https://github.com/bionetslab/ChemoTreeVsDL

## Ethics approval and consent to participate

The UKEr dataset used for this article is part of the PEDREF study and contains test results performed during patient care. Analysis of test results performed during patient care for research is in accordance with the applicable German and Bavarian regulations and does not require patients’ explicit consent. Use of pediatric and adult patient datasets in the PEDREF study has been approved by the Ethical Review Boards of the University Hospital Erlangen, reference number 97 17 Bc.

## Consent for publication

Not applicable.

## Author contributions

FR, CS, and DBB conceptualized the study and drafted the manuscript. FR and CS developed the software and carried out the analyses. JM, JZ, and MM provided critical feedback, with JZ and MM supporting the interpretation of clinical results. DBB supervised the work.

## Acknowledgements

The authors gratefully acknowledge the scientific support and HPC resources provided by the Erlangen National High Performance Computing Cen-ter (NHR@FAU) of the Friedrich-Alexander-Universitäat Erlangen-Nürnberg (FAU). The hardware is funded by the German Research Foundation (DFG).

## Supplementary information

Additional file 1 provides supporting material for the study: Table S1 lists the full set of medical codes and criteria used for cohort extraction in MIMIC–IV and UKEr. Figure S1 presents the AUC-PRCs and F1 scores of non-temporal models trained on all combinations of V, M, and D features. Figures S2–S4 report the feature importance results for the two MIMIC-IV cohorts and the aplasia UKEr cohort. Figure S5 reports the computational resource usage of all models.

## References

[1] Knevel R, Liao KP. From real-world electronic health record data to real-world results using artificial intelligence. Annals of the Rheumatic Diseases. 2023;82(3):306–311.

[2] Zhuang Q, Zhang AY, Tan RSYC, Yang GM, Neo PSH, Tan DSW, et al. Towards proactive palliative care in oncology: developing an explainable EHR-based machine learning model for mortality risk prediction. BMC Palliative Care. 2024;23(1):124.

[3] Lu SC, Xu C, Nguyen CH, Geng Y, Pfob A, Sidey-Gibbons C. Machine learning–based short-term mortality prediction models for patients with cancer using electronic health record data: systematic review and critical appraisal. JMIR Medical Informatics. 2022;10(3):e33182.

[4] Shickel B, Tighe PJ, Bihorac A, Rashidi P. Deep EHR: A survey of recent advances in deep learning techniques for electronic health record (EHR) analysis. IEEE Journal of Biomedical and Health Informatics. 2017;22(5):1589–1604.

[5] Gupta M, Gallamoza B, Cutrona N, Dhakal P, Poulain R, Beheshti R. An extensive data processing pipeline for MIMIC-IV. In: Proceedings of the Machine Learning for Health Conference. PMLR; 2022. p. 311–325.

[6] Reddy BK, Delen D. Predicting hospital readmission for lupus patients: an RNN-LSTM-based deep-learning methodology. Computers in Biology and Medicine. 2018;101:199–209.

[7] Rasheed N, Khaliq U, Ahmed A, Bermak A. Predicting patient ICU readmission using recurrent neural networks with long short-term memory. IEEE Access. 2025;.

[8] Men L, Ilk N, Tang X, Liu Y. Multi-disease prediction using LSTM recurrent neural networks. Expert Systems with Applications. 2021;177:114905.

[9] Krishnan S, Magalingam P, Ibrahim R. Hybrid deep learning model using recurrent neural network and gated recurrent unit for heart disease prediction. International Journal of Electrical & Computer Engineering (2088-8708). 2021;11(6).

[10] Che Z, Purushotham S, Cho K, Sontag D, Liu Y. Recurrent neural networks for multivariate time series with missing values. Scientific Reports. 2018;8(1):6085.

[11] Baytas IM, Xiao C, Zhang X, Wang F, Jain AK, Zhou J. Patient subtyping via time-aware LSTM networks. In: Proceedings of the 23rd ACM SIGKDD International Conference on Knowledge Discovery and Data Mining. New York, NY, USA: ACM; 2017. p. 65–74.

[12] Kidger P, Morrill J, Foster J, Lyons T. Neural controlled differential equations for irregular time series. In: Advances in Neural Information Processing Systems. vol. 33; 2020. p. 6696–6707.

[13] Tipirneni S, Reddy CK. Self-supervised transformer for sparse and irregularly sampled multivariate clinical time-series. ACM Transactions on Knowledge Discovery from Data (TKDD). 2022;16(6):1–17.

[14] Catling FJR, Wolff AH. Temporal convolutional networks allow early prediction of events in critical care. Journal of the American Medical Informatics Association. 2020;27(3):355–365.

[15] Bednarski BP, Singh AD, Zhang W, Jones WM, Naeim A, Ramezani R. Temporal convolutional networks and data rebalancing for clinical length of stay and mortality prediction. Scientific Reports. 2022;12(1):21247.

[16] Chen Yw, Li Yj, Deng P, Yang Zy, Zhong Kh, Zhang Lg, et al. Learning to predict in-hospital mortality risk in the intensive care unit with attention-based temporal convolution network. BMC Anesthesiology. 2022;22(1):119.

[17] Song H, Rajan D, Thiagarajan J, Spanias A. Attend and diagnose: Clinical time series analysis using attention models. In: Proceedings of the AAAI conference on artificial intelligence. vol. 32; 2018. .

[18] Jazayeri A, Liang OS, Yang CC. Imputation of missing data in electronic health records based on patients’ similarities. Journal of Healthcare Informatics Research. 2020;4(3):295–307.

[19] Getzen E, Ungar L, Mowery D, Jiang X, Long Q. Mining for equitable health: Assessing the impact of missing data in electronic health records. Journal of Biomedical Informatics. 2023;139:104269.

[20] Harutyunyan H, Khachatrian H, Kale DC, Ver Steeg G, Galstyan A. Multi-task learning and benchmarking with clinical time series data. Scientific Data. 2019;6(1):96.

[21] Iwase S, Nakada Ta, Shimada T, Oami T, Shimazui T, Takahashi N, et al. Prediction algorithm for ICU mortality and length of stay using machine learning. Scientific Reports. 2022;12(1):12912.

[22] Fridgeirsson EA, Sontag D, Rijnbeek P. Attention-based neural networks for clinical prediction modelling on electronic health records. BMC Medical Research Methodology. 2023;23(1):285.

[23] Sheehy J, Gallanagh M, Sullivan C, Lane S. Clinical prediction models for febrile neutropenia and its outcomes: a systematic review. Supportive Care in Cancer. 2025;33(7):537.

[24] Cho BJ, Kim KM, Bilegsaikhan SE, Suh YJ. Machine learning improves the prediction of febrile neutropenia in Korean inpatients undergoing chemotherapy for breast cancer. Scientific Reports. 2020;10(1):14803.

[25] Bozcuk H, Senol Coskun H, Ilhan Y, Göksu SS, Yıldız M, Bayram S, et al. Prospective external validation of an updated algorithm to quantify risk of febrile neutropenia in cancer patients after a cycle of chemotherapy. Supportive Care in Cancer. 2022;30(3):2621–2629.

[26] Wiberg H, Yu P, Montanaro P, Mather J, Birz S, Schneider M, et al. Prediction of neutropenic events in chemotherapy patients: A machine learning approach. JCO Clinical Cancer Informatics. 2021;5:904–911.

[27] Boccia R, Glaspy J, Crawford J, Aapro M. Chemotherapy-induced neutropenia and febrile neutropenia in the US: a beast of burden that needs to be tamed? The Oncologist. 2022;27(8):625–636.

[28] Klastersky J, De Naurois J, Rolston K, Rapoport B, Maschmeyer G, Aapro M, et al. Management of febrile neutropaenia: ESMO clinical practice guidelines. Annals of Oncology. 2016;27:v111–v118.

[29] Townsley DM, Dumitriu B, Young NS. Aplastic anemia: diagnosis and treatment. Mayo Clinic Proceedings. 2014;89(11):1645–1660.

[30] Johnson AE, Bulgarelli L, Shen L, Gayles A, Shammout A, Horng S, et al. MIMIC-IV, a freely accessible electronic health record dataset. Scientific data. 2023;10(1):1.

[31] Breiman L. Random forests. Machine learning. 2001;45(1):5–32.

[32] Friedman JH. Greedy function approximation: a gradient boosting machine. Annals of statistics. 2001;p. 1189–1232.

[33] Chen T, Guestrin C. Xgboost: A scalable tree boosting system. In: Proceedings of the 22nd acm sigkdd international conference on knowledge discovery and data mining; 2016. p. 785–794.

[34] Prokhorenkova L, Gusev G, Vorobev A, Dorogush AV, Gulin A. CatBoost: unbiased boosting with categorical features. Advances in Neural Information Processing Systems. 2018;31.

[35] Chung J, Gulcehre C, Cho K, Bengio Y. Gated feedback recurrent neural networks. In: International conference on machine learning. PMLR; 2015. p. 2067–2075.

[36] Hochreiter S, Schmidhuber J. Long short-term memory. Neural computation. 1997;9(8):1735–1780.

[37] Lea C, Flynn MD, Vidal R, Reiter A, Hager GD. Temporal convolutional networks for action segmentation and detection. In: proceedings of the IEEE Conference on Computer Vision and Pattern Recognition; 2017. p. 156–165.

[38] Shukla SN, Marlin B. Interpolation-Prediction Networks for Irregularly Sampled Time Series. In: International Conference on Learning Representations; 2019. .

[39] Bernett J, Blumenthal DB, Grimm DG, Haselbeck F, Joeres R, Kalinina OV, et al. Guiding questions to avoid data leakage in biological machine learning applications. Nat Methods. 2024;21(8):1444–1453.

[40] Johnson AEW, Pollard TJ, Shen L, Lehman LWH, Feng M, Ghassemi M, et al. MIMIC-III, a freely accessible critical care database. Sci Data. 2016;3(1):160035.

[41] Silva I, Moody G, Scott DJ, Celi LA, Mark RG. Predicting in-hospital mortality of ICU patients: The PhysioNet/computing in cardiology challenge 2012. Comput Cardiol (2010). 2012;39:245–248.

[42] Reyna MA, Josef CS, Jeter R, Shashikumar SP, Westover MB, Nemati S, et al. Early prediction of sepsis from clinical data: The PhysioNet/Computing in Cardiology Challenge 2019. Crit Care Med. 2020;48(2):210–217.

[43] Shukla SN, Marlin B. Multi-Time Attention Networks for Irregularly Sampled Time Series. In: International Conference on Learning Representations; 2021. .

[44] Li Z, Li S, Yan X. Time series as images: Vision transformer for irregularly sampled time series. Advances in Neural Information Processing Systems. 2023;36:49187–49204.

[45] Liu J, Cao M, Chen S. Musicnet: A gradual coarse-to-fine framework for irregularly sampled multivariate time series analysis. arXiv preprint arXiv:241201063. 2024;.

[46] Huang J, Yang B, Yin K, Xu J. Dna-t: Deformable neighborhood attention transformer for irregular medical time series. IEEE Journal of Biomedical and Health Informatics. 2024;28(7):4224–4237.

[47] Yu Z, Xu C, Jin Y, Wang Y, Zhao J. Smart: Towards pre-trained missing-aware model for patient health status prediction. Advances in Neural Information Processing Systems. 2024;37:63986–64009.

[48] Li X, Teng F, Ma M, Qin Z, Guo J, Xu J, et al. Multi-level transfer learning for irregular clinical time series prediction. Knowledge-Based Systems. 2025;p. 113825.

[49] Liu J, Cao M, Chen S. Timecheat: A channel harmony strategy for irregularly sampled multivariate time series analysis. In: Proceedings of the AAAI Conference on Artificial Intelligence. vol. 39; 2025. p. 18861–18869.

[50] Liu J, Cao M, Chen S. Beyond Observations: Reconstruction Error-Guided Irregularly Sampled Time Series Representation Learning. arXiv preprint arXiv:251106854. 2025;.

[51] Chowdhury RR, Li J, Zhang X, Hong D, Gupta RK, Shang J. Primenet: Pre-training for irregular multivariate time series. In: Proceedings of the AAAI Conference on Artificial Intelligence. vol. 37; 2023. p. 7184–7192.

52. Patel H, Qiu R, Irwin A, Sadiq S, Wang S. EMIT-Event-Based Masked Auto Encoding for Irregular Time Series. In: 2024 IEEE International Conference on Data Mining (ICDM). IEEE; 2024. p. 370–379.

[53] Restrepo D, Wu C, Jia Y, Sun JK, Gallifant J, Bielick CG, et al. Representation learning of lab values via masked autoencoder. arXiv preprint arXiv:250102648. 2025;.

[54] Grinsztajn L, Oyallon E, Varoquaux G. Why do tree-based models still out-perform deep learning on typical tabular data? Advances in Neural Information Processing Systems. 2022;35:507–520.

[55] McElfresh D, Khandagale S, Valverde J, Prasad C V, Ramakrishnan G, Goldblum M, et al. When do neural nets outperform boosted trees on tabular data? Advances in Neural Information Processing Systems. 2023;36:76336–76369.

[56] Zhang J, Zheng S, Cao W, Bian J, Li J. Warpformer: A multi-scale modeling approach for irregular clinical time series. In: Proceedings of the 29th ACM SIGKDD Conference on Knowledge Discovery and Data Mining; 2023. p. 3273–3285.

