## additional file 1 for "Non-temporal tree-based models outperform temporal deep learning models in the prediction of chemotherapy-induced side effects from longitudinal laboratory data"

Farnaz Rahimi†, Christel Sirocchi†, Julian Matschinske,
Markus Metzler, Jakob Zierk, David B. Blumenthal

†These authors contributed equally to this work.

**Table S1**: ICD-10 codes, OPS codes, and criteria used for cohort extraction from the MIMIC-IV and UKEr datasets.

| | **MIMIC-IV** | **UKEr** |
|---|---|---|
| **Cancer diagnosis** | ICD-10 diagnosis code C* | |
| **Chemotherapy application** | ICD-10 diagnosis code Z5111 **OR** one of the following ICD-10 procedure codes for introduction of antineoplastic into vein, artery, subcutaneous tissue, spinal canal, or epidural space: 3E03005, 3E0300M, 3E03305, 3E0330M, 3E04005, 3E0400M, 3E04305, 3E0430M, XW03336, XW03351, XW03358, XW033B3, XW033D6, XW033Q5, XW033S5, XW04336, XW04351, XW04358, XW043B3, XW043D6, XW043Q5, XW043S5, 3E05005, 3E0500M, 3E05305, 3E0530M, 3E06005, 3E0600M, 3E06305, 3E0630M, 3E01305, 3E0130M, XW01348, 3E0D305, 3E0D30M, 3E0D705, 3E0D70M, 3E0DX05, 3E0DX0M, XW0DXJ5, XW0DXL5, XW0DXR5, XW0DXV5, 3E0R305, 3E0R30M, 3E0S305, 3E0S30M | ICD-10 diagnosis code Z511 or Z512 **OR** one of the following OPS procedure codes: 8-542*, 8-543*, 8-544* |
| **Aplasia** | Item ID 52075 < 0.5 (neutrophil count) **OR** one of the following ICD-10 procedure codes for transfusion of red blood cells or platelets: 30230N0, 30230N1, 30230R0, 30230R1, 30233N0, 30233N1, 30233R0, 30233R1, 30240N0, 30240N1, 30240R0, 30240R1, 30243N0, 30243N1, 30243R0, 30243R1, 30250N0, 30250N1, 30250R0, 30250R1, 30253N0, 30253N1, 30253R0, 30253R1, 30260N0, 30260N1, 30260R0, 30260R1, 30263N0, 30263N1, 30263R0, 30263R1, 30273N1, 30273R1, 30277N1, 30277R1, 302A3N0, 302A3N1, 302A3R0, 302A3R1 | LOINC code 751-8 < 0.5 (neutrophil count) **OR** OPS procedure code 8-800* (transfusion) |
| **Neutropenic fever** | ICD-10 diagnosis code D70* (neutropenia) **AND** ICD-10 diagnosis code R50* (fever) | |

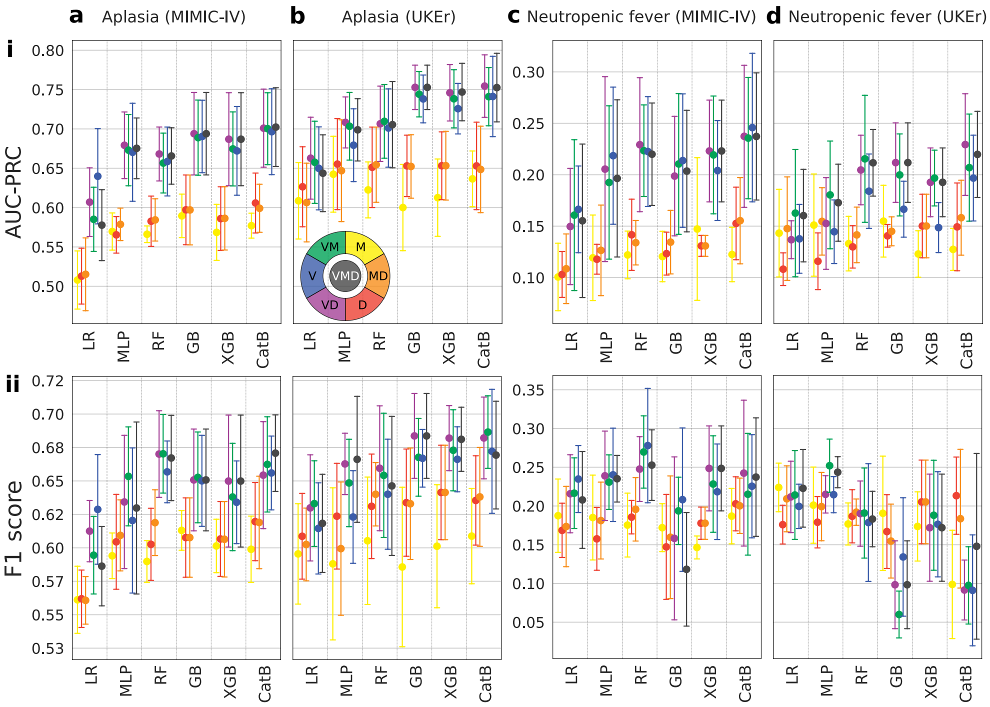


**Fig. S1**: Performance of non-temporal models trained on all combinations of V (discretised imputed values), M (missingness indicator), and D (temporal gap) features across four datasets: the aplasia cohort from MIMIC-IV (a) and UKEr (b), and the neutropenic fever cohort from MIMIC-IV (c) and UKEr (d). Plots show mean ± standard deviation of test AUC-PRC (i) and F1 scores (ii) computed on the outer folds of a nested 5×3 CV.

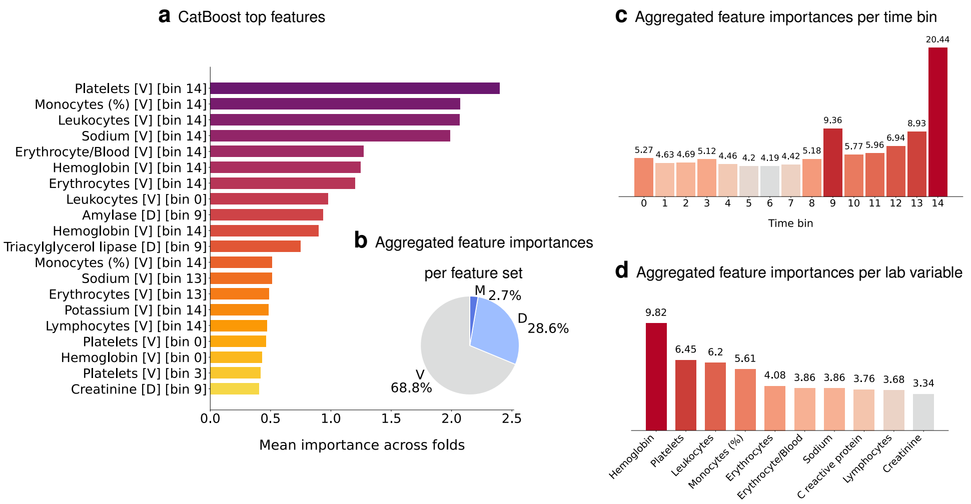


**Fig. S2**: Analysis of CatBoost feature importance scores for the aplasia cohort from the UKEr dataset (V/M/D features, daily resolution, 5-fold CV). (b–d) Contributions of feature sets (b), time bins (c), and laboratory variables (d) assessed by summing the importance scores for each feature type within its respective category.

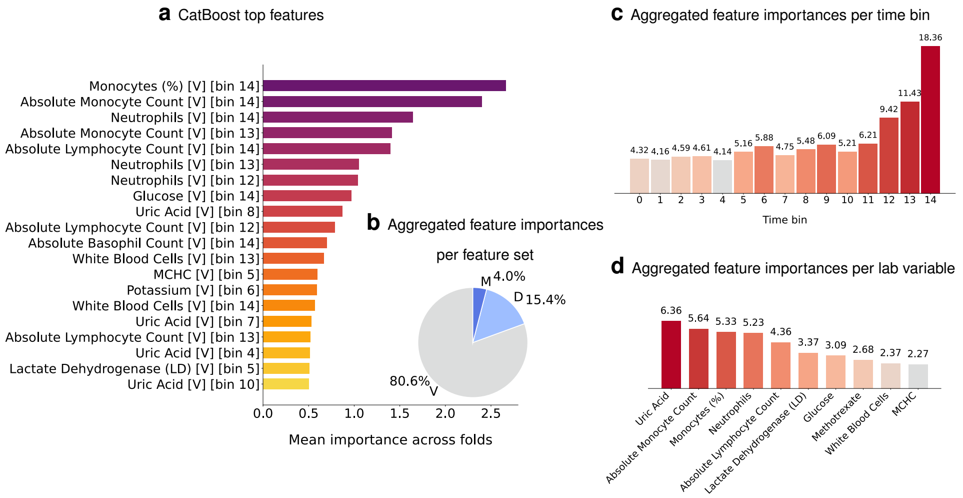


**Fig. S3**: Analysis of CatBoost feature importance scores for the neutropenic fever cohort from the MIMIC-IV dataset (V/M/D features, daily resolution, 5-fold CV). (b–d) Contributions of feature sets (b), time bins (c), and laboratory variables (d) assessed by summing the importance scores for each feature type within its respective category.

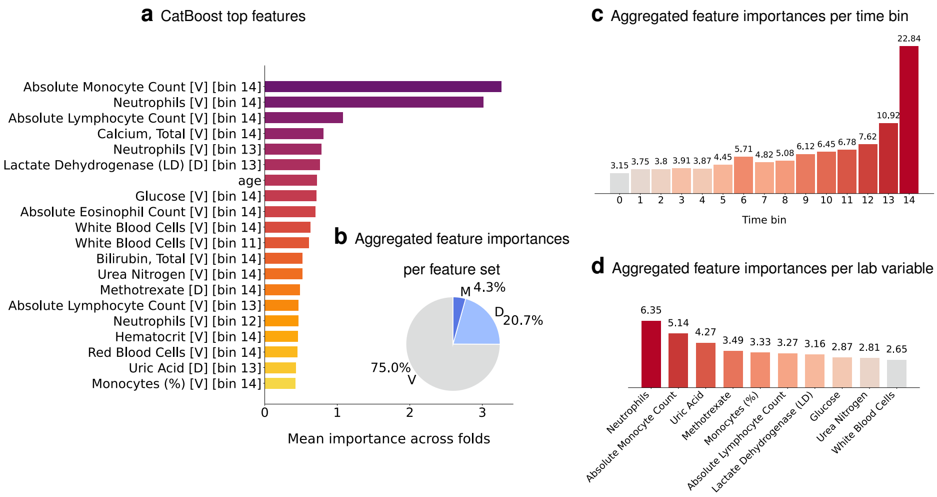


**Fig. S4**: Analysis of CatBoost feature importance scores for the aplasia cohort from the MIMIC-IV dataset (V/M/D features, daily resolution, 5-fold CV). (b–d) Contributions of feature sets (b), time bins (c), and laboratory variables (d) assessed by summing the importance scores for each feature type within its respective category.

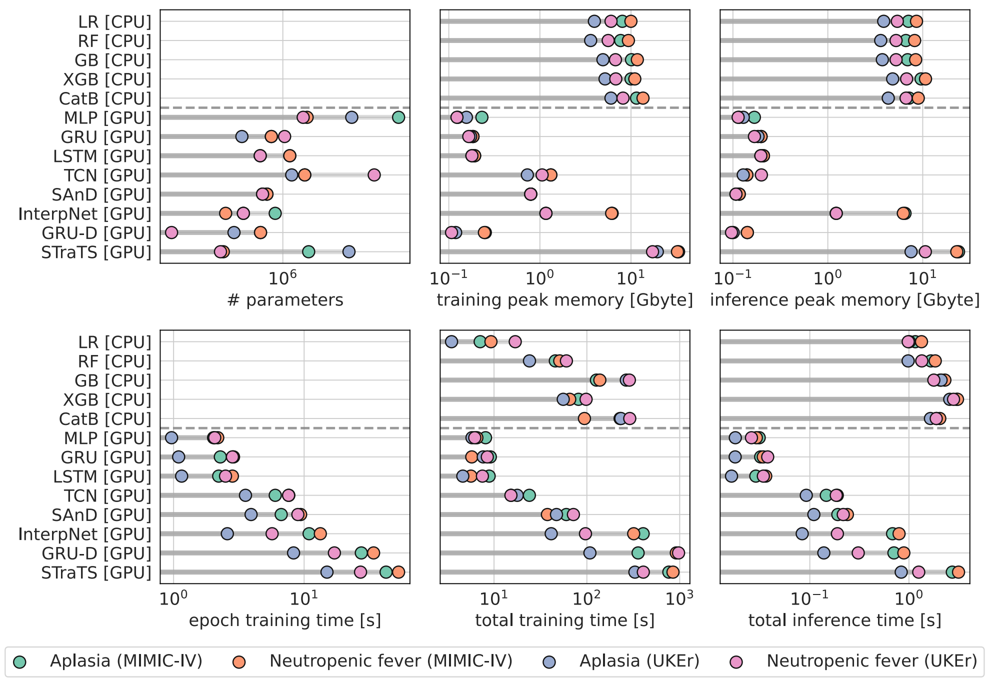


**Fig. S5**: Computational resource usage for all models, measured as training and inference time and memory usage, and total number of parameters for neural networks.
